# Stethoscope and non-infrared thermometer disinfection among physicians: A cross-sectional study with implications for the control of COVID-19

**DOI:** 10.1101/2020.08.14.20174433

**Authors:** Biniyam Sahiledengle, Yohannes Tekalegn, Kebebe Bekele, Abdi Tesemma, Bruce John Edward Quisido

**Author notes:** Correspondence: Biniyam Sahiledengle, Public Health Department, Madda Walabu University Goba Referral hospital, School of Health Sciences, Bale-Goba, Ethiopia, P.o.box: 76.

## Abstract

**Background:** Stethoscopes and non-infrared thermometers are the customary medical equipment used by the physicians on a daily basis, among various patients. With the rise of potential infections in the healthcare facilities and the transmission nature of the current COVID-19 pandemic, consistent and correct disinfections of these devices after each use should not be pardoned. This study, therefore, describes the level of stethoscope and non-infrared thermometer disinfection practices among physicians working in healthcare facilities during the COVID-19 pandemic.

**Methods:** An online survey was circulated using an anonymous and self-reporting questionnaire via Google form with a consent form appended to it.

**Results:** The proportion of stethoscope and non-Infrared thermometer disinfections after every use was 13.9% (95%CI: 10.9-17.6) and 20.4% (95%CI: 16.7-24.5), respectively. Taking COVID-19 training (AOR: 2.52; 95%CI: 1.29-4.92) and the availability of stethoscope disinfection materials at the workplace (AOR: 3.03; 95% CI: 1.29-7.10) were significantly increased the odds of stethoscope disinfection after every use. The odds of stethoscope disinfection after every use was significantly decreased for those who reported the use of shared stethoscope (AOR: 0.34; 95% CI: 0.12-0.92).

**Conclusion:** Only a wee share of the respondents reported that they have disinfected their stethoscopes and non-infrared thermometers after every use – possibly jeopardizing both patients and clinicians safety, particularly during the COVID-19 pandemic.

## BACKGROUND

Stethoscopes are the most customary medical equipment wield by physicians on a daily basis, among various patients. With the rise of potential infections in the healthcare setups, stethoscope disinfection should not be pardoned [1-4]. The Gold Standard is to disinfect stethoscopes after each contact with patients [5]. Current studies suggest that disinfecting stethoscopes between each patient contact using a 70% isopropyl alcohol swab or alcohol pad from the bell to the earpieces, including the tubing, is effective in eradicating bacteria including methicillin-resistant *Staphylococcus aureus* [6,7]. However, only a minority of healthcare workers regularly disinfect their stethoscopes [2,8,9].

It is well-documented that the stethoscopes can harbor pathogenic microorganisms [3,10-22]. Potential pathogens cultured from stethoscopes include *Pseudomonas aeruginosa* [3,9], *Clostridium Difficile* [11], methicillin-resistant *Staphylococcus aureus (MRSA)* [3,13-16], Vancomycin-resistant *enterococci* [3,18,19], and *Acinetobacter baumannii* [20]. Huang et al reported that highly resistant bacteria, MRSA can potentially linger up to 9 days on stethoscopes [23]. Additionally, a review by Wolfensberger et al found that the diaphragms of the stethoscopes become colonized by bacteria quickly; acquiring more pathogens than any part of the doctors’ hands except the fingertips [24].

Assessing patients’ body temperatures are one of the vital procedures in monitoring health conditions. In practice, patients’ body temperatures are taken at least twice daily among hospitalized patients, and the common anatomical sites for measurements are oral, rectal, and axillary temperatures. In low-income settings, acquiring patients’ body temperatures using non-infrared thermometers are still predominant practices, and these devices travel among healthcare workers across the hospital. This entails that non-infrared thermometers are often exposed to body fluids, and travel without proper disinfection may advance the spread of cross-infection [25-27]. Rectal thermometers and reusable oral thermometers may possibly contact with body fluids, hence, wager contaminations, and therefore are unsuitable in this travel fast COVID-19 pandemic situation [28]. Currently, non-contact infrared thermometer scans are preferable as they don’t require direct contact with the skin, and thus, lead to efficiency, expediency, and lessening the risks of cross-contaminations.

The Center for Disease Control and Prevention (CDC) classify stethoscope as both non-critical and semi-critical medical devices – depending on the association it has with intact and non-intact skins [29]. To this day, the role of stethoscopes as disease transmitters have been debated, and ultimately, these medical devices should be considered risks as it comes into contact with both patients and healthcare providers [3, 30,31]. The analyses of 22 studies reveal that Severe Acute Respiratory Syndrome (SARS) coronavirus, Middle East Respiratory Syndrome (MERS) coronavirus, or endemic human coronaviruses (HCoV) persist on inanimate surfaces like metals, glasses or plastics for up to 9 days [32]. This poses concerns among patients contracting COVID-19 from contaminated stethoscopes and non-infrared thermometers, even if this hasn’t been directly documented yet – at least the theoretical risk of COVID-19 is anticipated to be existing, and just as same as how one cleans his hands regularly, cleaning anything that contacts the patients would be potentially beneficial to their safety – as evidence suggests that human-to-human transmission of novel human coronavirus and were explicated with incubation times, and its spread via droplets, contaminated hands or objects (such as medical equipments) [33].

In Ethiopia, physicians habitually carry their stethoscopes around their necks or in their pouches, and on occasions brings them to their homes, as well. These consequently augments the risk of transmission of infections from the hospitals to homes, and vice versa. Up to date, no specific guidelines have been available in the country solely on the disinfection of this medical equipment [34]. To our knowledge, there are no available studies on this aspect that determine the level of physician’s stethoscopes and non-infrared thermometer disinfection at the national level. Consequently, the assessment of the current disinfection practices among physicians is essential. This study, therefore, describes the level of stethoscope and non-infrared thermometer disinfection practices among physicians and determine its associated factors during the COVID-19 pandemic.

## METHODS

### Study Design and Setting

This was a cross-sectional online survey conducted in Ethiopia from June 1 to 20, 2020 using Google form with a consent form appended to it. An internet-based survey was utilized due to the current COVID-19 pandemic and the government’s strict regulations on face-to-face interviews in Ethiopia. Ethiopia is located in the horn of Africa. It is one in the world with low medical doctor densities (0.769/10,000 population), which is far below the minimum threshold density. The number of physicians increased to 8,395 (in the year 2018) from a number of 1,936 in 2003. Withal, there are 2,528 specialist medical practitioners in Ethiopia [35].

### Study Participants and Eligibility

All medical – doctors, specialist medical practitioners, residents, and interns – who are able to access and utilize at least an Email, Facebook, LinkedIn, Telegram, and by Tweeter, who consented to participate, and those working in healthcare facilities were eligible in the survey. With exceptions, physicians who do not engage in direct contact with patients and those works in any administrative areas were excluded.

### Sample Size Determination

The sample size was determined using Epi Info™ 7.1.1.14 statistical software (Center for Disease Control and Prevention, 2013) using single population proportion formula with the assumption of 95% confidence level, 5% precision, considering the proportion of healthcare workers who had safe stethoscope disinfection practice was 39.7% [2], and considering 25% non-response rate. The calculated sample size was (n=460).

### Data Collection

Data were collected via self-administered structured questionnaires. The study participants who were willing to partake in the present survey and could access the Google form link obtain an informed consent sheet as a pre-requirement before proceeding to participate in the actual survey. Explicit information about the aim, scope, and eligibility criteria along with the link was purveyed. The call for participation was dispatched thru Emails, Messenger, Tweeter, Linked In, and Telegram. Upon receiving the Google form link, the participants were auto directed to the information about informed consent and their voluntariness to participate.

### Instrument

An internet-based, self-administered closed-ended questionnaire was utilized. The online data collection tools were created using the Google Forms provided by Google ™ and were constructed using the English language. The tools consisted of four sections, with a total of 25 questions. The first section was analogous to the participants’ socio-demographic characteristics – including age, gender, professional titles, working department, service year, type of healthcare facility, place of residence, and history of COVID-19-related training. The second section comprised of six questions relating to stethoscope disinfection practices during COVID-19 pandemic (such as “Have you ever disinfected your stethoscope?”, “How often do you disinfect your stethoscope?”, “Which stethoscope part you frequently disinfect?”, and “Do you disinfect your stethoscope after examining your last patient?” along with questions about reasons for not disinfecting their stethoscopes. The third section of the questionnaire encompassed four questions pertaining to topics about non-infrared thermometer disinfections. Finally, the last section composed of five questions about the physicians’ awareness related to stethoscope and non-infrared thermometer disinfections. The data collection tool was tested for internal consistency and the resulting Cronbach’s Alpha value of 0.841 was obtained.

### Study Variables and Measurements

#### Dependent Variables

In the physicians’ self-report regarding stethoscope and non-infrared thermometer disinfection practices, respondents were posed to specify their usual practices (during COVID-19 pandemic) by uttering to them “How often do you disinfect your stethoscope?”. Physicians who claimed that they have disinfected their stethoscopes after every use/after contact with patients were coded as “1” and labeled as “disinfection after every use”, if otherwise “0”. Similarly, physicians who have claimed that they have disinfected non-infrared thermometers after every use were coded as “1”, and zero, if otherwise.

#### Independent Variables

The independent variables include gender, age, years of service, type of healthcare facility, availability of reminders about medical equipment disinfections in their respective working units (e.g: poster), availability of disinfectant solutions in their working units, and awareness on stethoscope and non-infrared thermometer disinfections.

#### Data Analysis

Completed questionnaires were extracted from Google Forms in Excel spreads and were exported to STATA version 14.0 for the analyses. Descriptive statistics were employed to illustrate the data. Multiple binary logistic regression models were used to assess factors associated with the outcome variables (stethoscope and non-infrared thermometer disinfection after every use). All the independent variables were tested for potential multicollinearities before placing them in the multivariable logistic regression models. The multicollinearity effects were assessed with a cut of off point of variation inflation factor (VIF) of greater than ten. Finally, significant variables were discerned based on the adjusted odds ratio (AOR) with 95% Confidence Intervals (CIs), and results were deemed significant if they reflect p< 0.05. To scrutinize the accuracy of the final formulated model, the Hosmer-Lemeshow test for the overall goodness of fit was used. Accordingly, the overall goodness of fit was 0.799 and 0.163 for stethoscope and non-infrared thermometer disinfection after every use models, respectively.

#### Ethical approval and considerations

The study was conducted according to the Declaration of Helsinki. Ethical approval was granted by the Institutional Review Board (IRB) at Madda Walabu University. The respondents penned the online questionnaires anonymously, voluntarily, and independently. The privacy and confidentiality of the study participants were also safeguarded throughout the data collection. An information letter was incorporated on the first page of every questionnaire which covered information about the study description, eligibility criteria, voluntary participation, and confidentiality. Also, the contact details of the investigators (name, phone number, email address, and their affiliations) for any inquiry were mounted on the top of the questionnaire. All participants were assumed to only proceed on the survey after reading the consent and acknowledging engagement. Accordingly, electronic informed consent was sought from the study participants as pre-requisites before they are able to annex the survey.

## RESULTS

### Socio-Demographic Characteristics of the Study Participants

A total of 422 (response rate of 91.7%) physicians filled the online forms. The study participants composed – 62.8% medical doctors, 33.6% specialist medical practitioners, and 3.6% residents. Of these, 368 (87.20%) were males, 388 (91.94%) are currently working in governmental healthcare facilities, and 238 (56.40%) have less than 3 years of work experience. The mean age of the study participants was 30.13 (± 4.34) years (range: 22-55) and 228 (54.03%) were in the age group of 26-30 (**Table I**).

**Table I:** Socio-Demographic Characteristics of the Study Participants.

| **Variables** | **Category** | **n** | **%** |
| --- | --- | --- | --- |
| Profession | Medical doctor <sup>a</sup> | 265 | 62.80 |
|  | Specialist medical practitioners | 142 | 33.65 |
|  | Resident physician | 15 | 3.55 |
| Gender | Male | 368 | 87.20 |
|  | Female | 54 | 12.80 |
| Age | 20-25 | 44 | 10.43 |
|  | 26-30 | 228 | 54.03 |
|  | 31-35 | 107 | 25.36 |
|  | 36-40 | 35 | 8.29 |
|  | 40+ | 8 | 1.90 |
| Service year | 1-3 | 238 | 56.40 |
|  | 4-6 | 97 | 22.90 |
|  | 7-9 | 37 | 8.77 |
|  | 10+ | 50 | 11.85 |
| Training on COVID-19 | Yes | 107 | 25.36 |
|  | No | 315 | 74.64 |
| Current working healthcare facility | Governmental | 388 | 91.94 |
|  | Private | 35 | 8.06 |
| Department | Surgical | 113 | 26.78 |
|  | Outpatient Department (OPD) | 106 | 25.12 |
|  | Medical | 47 | 11.14 |
|  | Gynecology and Obstetrics Department | 37 | 8.77 |
|  | Operation Room (OR) | 32 | 7.58 |
|  | Emergency (E-OPD) | 31 | 7.35 |
|  | Pediatric Department | 29 | 6.87 |
|  | COVID-19 Center | 14 | 3.32 |
|  | Intensive Care Unit (ICU) | 6 | 1.42 |
|  | Pathology Department | 2 | 0.47 |
|  | Radiology Department | 2 | 0.47 |
|  | Psychiatric Department | 1 | 0.24 |
|  | Orthopedic Department | 1 | 0.24 |
|  | Urology Department | 1 | 0.24 |

<sup>a</sup>including medical interns

### Stethoscope Disinfection

As seen in **Table II**, the proportion of stethoscope disinfection after every use was 13.9% (95%CI: 10.9-17.6). A quarter of the physicians delineated that they never disinfected their stethoscopes. Fifty-four percent of them also reported spending only 20 or fewer seconds per stethoscope disinfection. The proportion of the physicians who reported about disinfecting the different parts of the stethoscope variegated as well (diaphragm, 39.51%; diaphragm, tubing, and earpieces, 28.40%; diaphragm and earpieces, 23.77%, earpieces, 7.72%; plastic tubing, 0.62%). **Figure 1** exhibited the reported barriers to stethoscope disinfection. More than one third (35.9%) of the physicians outlined that they cried out for access to stethoscope disinfectants when needed. Factors, such as forgetfulness (29.3%) and negligence (25.2%) were the frequently cited barriers to such. Nevertheless, 378(89.57%) of respondents believed that stethoscopes necessitated to be disinfected after every use.

**Figure 1:**
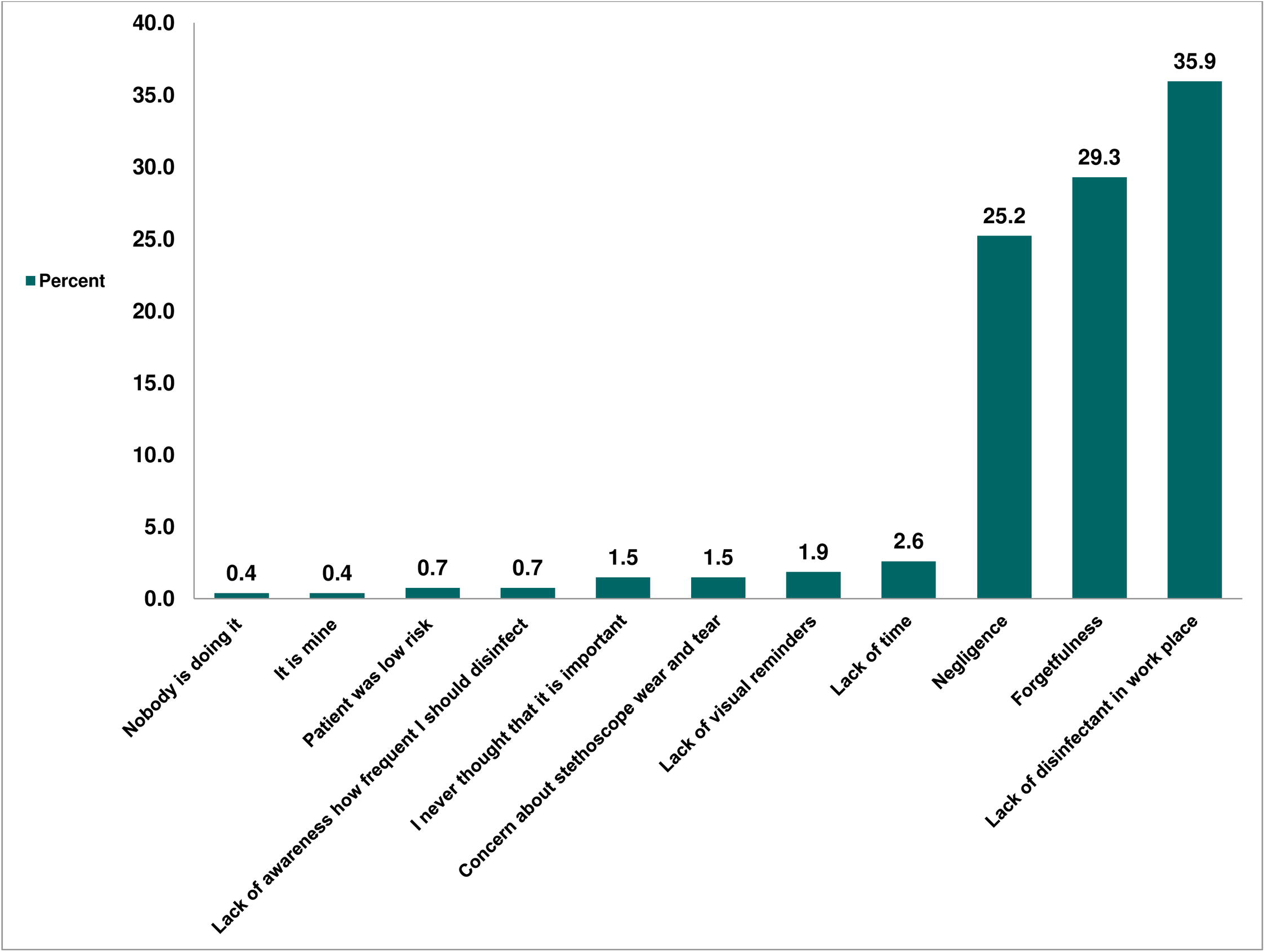
Bar graph showing percentage of reported obstacles to stethoscope disinfection among physicians during COVID-19

#### Factors associated with stethoscope disinfection after every use

The odds ratios for a unit change on each covariate are delineated thru the binary logistic regression model in **Table III**. On fixed values of the other covariates, young physicians (age ≤ 30) nearly have three times more the odds of disinfection after every use than those of age greater than 30 (adjusted odds ratio [AOR], 2.52; 95% confidence interval [CI]: 1.10-5.78). Those who have had previous COVID-19 training preceding the survey had approximately three times favorable odds of disinfection after every use than those who have none (AOR: 2.52; 95% CI: 1.29-4.92). In addition, the availability of stethoscope disinfectants at the workplace significantly increased the odds of disinfection after every use (AOR: 3.03; 95% CI: 1.29-7.10). Amid the participants, disinfection after every use was significantly greater among resident physicians compared to those of general practitioners (AOR: 4.61; 95%CI: 1.29-16.52). On the contrary, the odds of disinfection after every use were significantly lesser among those who reported using a shared stethoscope (AOR: 0.34; 95% CI: 0.12-0.92).

**Table II:** Stethoscopes Disinfection Practices and Physicians Perceptions.

| **Variables** | **Category** | **n** | **%** |
| --- | --- | --- | --- |
| Stethoscope ownership | Yes/my own | 340 | 80.57 |
|  | Yes/shared | 82 | 19.43 |
| How often do you disinfect your stethoscope ?* | After every use | 59 | 13.98 |
|  | Monthly | 13 | 3.08 |
|  | Multiple times per day not after every use | 26 | 6.16 |
|  | Multiple times per week but not every day | 34 | 8.06 |
|  | Never disinfect | 108 | 25.59 |
|  | Once a week or less often | 42 | 9.95 |
|  | Once or twice a day | 29 | 6.87 |
|  | Only after seeing high-risk patients | 111 | 26.30 |
| Disinfected stethoscope after the last patient? | Yes | 121 | 28.67 |
|  | No | 254 | 60.19 |
|  | I don't remember | 47 | 11.14 |
| Which part/s of the stethoscope do you frequently disinfect? (n=324) | Diaphragm | 128 | 39.51 |
|  | Diaphragm, tubing, and earpieces | 92 | 28.40 |
|  | Diaphragm and earpieces | 77 | 23.77 |
|  | Earpieces | 25 | 7.72 |
|  | Plastic tubing | 2 | 0.62 |
| Average duration of stethoscope disinfection (n=273) ,<br>Mean (SD): 22.76 (± 21.46)<br>Range: (2-120 Second) | <20 | 142 | 52.01 |
|  | ≥ 20 | 131 | 47.99 |
| Stethoscopes used on intact skin should be disinfected between each patient? | Yes | 70 | 16.59 |
|  | No | 352 | 83.41 |
| As in the case of skin that is not intact (e.g., trauma), stethoscopes should be disinfected before use on each patient? | Agree | 411 | 97.39 |
|  | Disagree | 3 | 0.71 |
|  | Not sure | 8 | 1.90 |
| Do stethoscopes need to be disinfected after every use? | Agree | 378 | 89.57 |
|  | Disagree | 18 | 4.27 |
|  | Not sure | 26 | 6.16 |
| Stethoscopes can contribute to the transmission of COVID-19 if not disinfected after every use? | Agree | 384 | 91.00 |
|  | Disagree | 1 | 0.24 |
|  | Not sure | 37 | 8.77 |
| Are there any reminders about medical equipment disinfection in your working unit? (e.g. poster) | Yes | 70 | 16.59 |
|  | No | 352 | 83.41 |
| Do you have a disinfectant solution in your working unit ? ( e.g. alcohol, alcohol-based sanitizer,...) | Yes | 302 | 71.56 |
|  | No | 120 | 28.44 |
| How often are you involved in direct contact with a known case of COVID-19 patient? (n=411) | Daily | 91 | 21.56 |
|  | Never/ not yet | 331 | 78.44 |

♣ Practice of disinfection was specific time during the COVID-19 pandemic.

**Table III:** Factors Associated with Stethoscope Disinfection among Physicians.

| Variables | Category | Disinfect after every use (%) | COR (95%CI) | p-value | AOR (95%CI) |
| --- | --- | --- | --- | --- | --- |
| Gender | Male | 56 (94.9) | 1 |  | 1 |
|  | Female | 3 (5.1) | 0.32(0.09-1.08) | 0.06 | 0.32(0.09-1.14) |
| Age | ≤30 | 47 (79.7) | 2.40(1.23-4.68)* | 0.01 | 2.52(1.10-5.78)** |
|  | >30 | 12 (20.3) | 1 |  | 1 |
| Service year | 1-3 | 32 (54.2) | 1 |  |  |
|  | 3+ | 27 (45.8) | 1.12(0.63-1.92) | 0.72 |  |
| Training on COVID-19 | Yes | 24 (40.7) | 2.31(1.30-4.11)* | 0.004 | 2.52(1.29-4.92)** |
|  | No | 35 (59.3) | 1 |  | 1 |
| Profession | Medical doctor | 38 (64.4) | 1 |  | 1 |
|  | Specialist medical practitioners | 15 (25.4) | 0.70(0.37-1.33) | 0.28 | 0.97(0.41-2.31) |
|  | Resident physician | 6 (10.2) | 3.98(1.34-11.82)* | 0.01 | 4.61(1.29-16.52)** |
| Current working healthcare facility | Governmental | 57 (96.6) | 2.75(0.64-11.82) | 0.17 | 2.24(0.47-10.62) |
|  | Private | 2 (3.4) | 1 |  | 1 |
| Department | OPD & Emergency | 17 (28.8) | 1 |  | 1 |
|  | Surgical Department & OR | 19 (32.2) | 1.06(0.53-2.14) | 0.86 | 1.01(0.42-2.46) |
|  | Medical Department | 4 (6.8) | 0.66(0.21-2.06) | 0.47 | 0.57(0.17-1.94) |
|  | Gynecology and Obstetrics Department | 6 (10.2) | 1.36(0.49-3.75) | 0.54 | 1.27(0.38-4.26) |
|  | Pediatric Department | 5 (8.5) | 1.47(0.49-4.37) | 0.48 | 1.02(0.29-3.52) |
|  | COVID-19 Center | 7 (11.9) | 7.05(2.20-22.61)* | 0.001 | 3.11(0.84-11.50) |
|  | Intensive Care Unit (ICU) & others | 1 (1.7) | 0.59(0.07-4.81) | 0.62 | 0.32(0.03-3.12) |
| Ownership of stethoscope | My own | 54 (91.5) | 1 |  | 1 |
|  | Shared | 5 (8.5) | 0.34(0.13-0.89)* | 0.02 | 0.34(0.12-0.92)** |
| Availability of disinfectant in the work place | Yes | 50 (84.7) | 2.44(1.16-5.14)* | 0.01 | 3.03(1.29-7.10)** |
|  | No | 9 (15.3) | 1 |  | 1 |
| Are there any reminders about medical equipment disinfection in your working unit? (e.g. poster) | Yes | 17 (28.8) | 2.37 (1.25-4.46)* | 0.008 | 1.62(0.78-3.36) |
|  | No | 42 (71.2) | 1 |  | 1 |
| Stethoscopes used on intact skin should be disinfected between each patient? | Yes | 57 (96.6) |  |  |  |
|  | No | 2 (3.4) |  |  |  |
| Duration of stethoscope disinfection | <20 seconds | 26 (52.0) | 0.99(0.54-1.84) | 0.99 |  |
|  | ≥ 20 seconds | 24 (48.0) | 1 |  |  |
| How often are you involved in direct contact with a known case of COVID-19 patient? | Daily | 20 (33.9) | 2.10(1.16-3.83)* | 0.01 | 1.22(0.60-2.47) |
|  | Never/not yet | 39 (66.1) | 1 |  | 1 |

Hosmer-Lemeshow = 0.7992; Crude odds ratio: COR ; Adjusted odds ratio: AOR; \*p-value < 0.05 (Crude); \*\*p-value < 0.05 (Adjusted)

#### Non-Infrared Thermometer Disinfection

Three hundred sixty-six respondents, 86.73% concurred that the non-Infrared thermometers need to be disinfected after every use, to which 20.37% (95%CI: 16.78-24.51) reported on disinfecting succeeding every use, and two fifths (40.53%) of physicians sadly reported that they have never disinfected any thermometers (**Figure 2**). In this study, 141 (33.41%) of the respondents reported that they have disinfected the non-Infrared thermometers the last time they used them (**Table A.1**).

**Figure 2:**
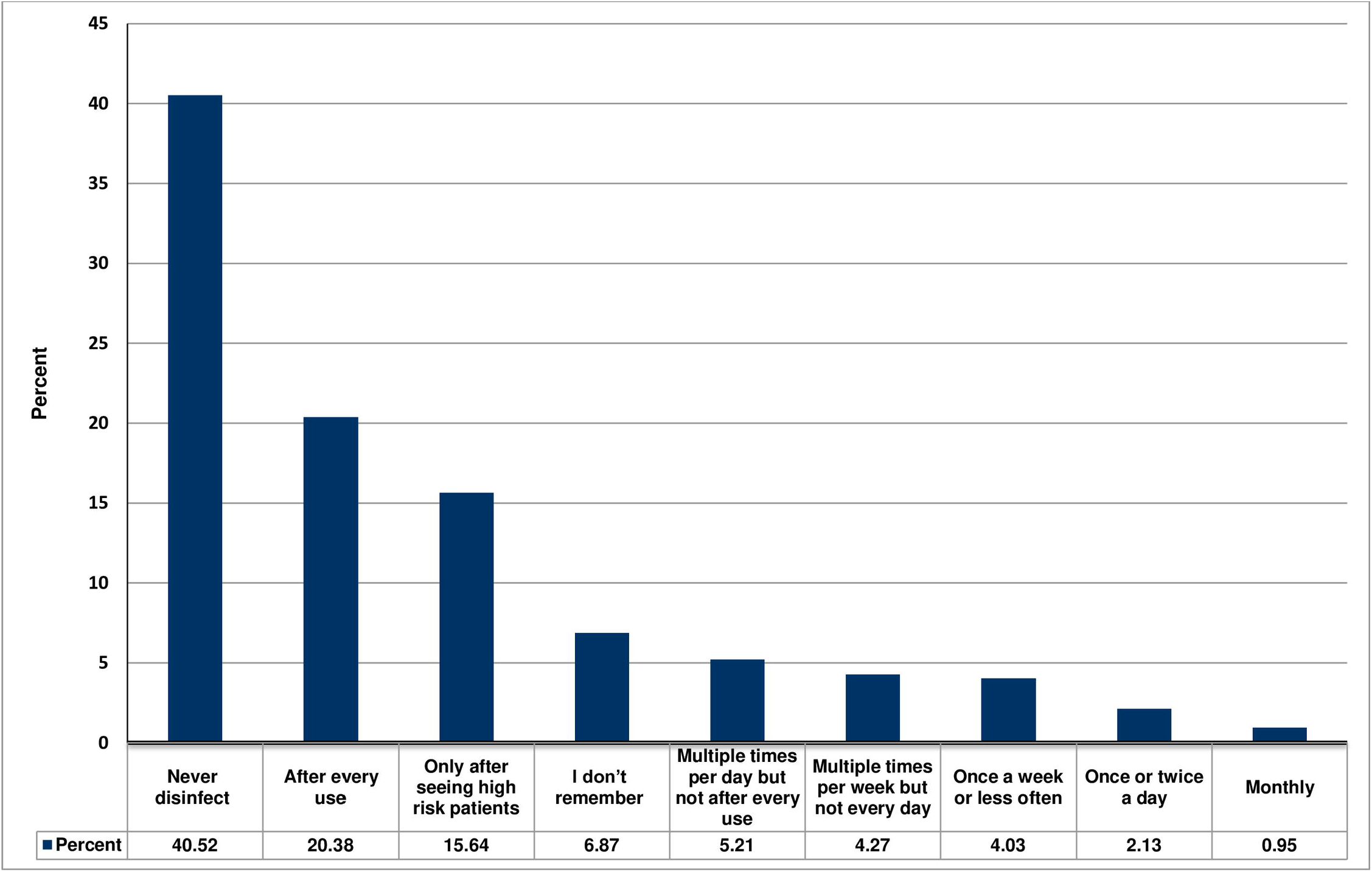
Bar graph showing percentage of non-Infrared thermometer disinfection during COVID-19 during COVID-19

#### Factors Associated With Non-Infrared Thermometer Disinfection after Every Use

In multivariable analyses, the odds of disinfection after every use were significantly higher among female physicians as compared to the males (AOR: 2.21; 95%CI: 1.21-4.36). Among the physicians, the odds of disinfection after every use were significantly higher in resident physicians (AOR: 7.10; 95% CI: 2.30-21.95) than those of general practitioners (**Table IV**).

**Table IV:** Factors Associated with Non-Infrared Thermometer Disinfection.

| Variables | Category | Disinfect after every use (%) | COR (95%CI) | p-value | AOR (95%CI) |
| --- | --- | --- | --- | --- | --- |
| Gender | Male | 69 (80.2) | 1 |  | 1 |
|  | Female | 17 (19.8) | 1.99(1.06-3.74)* | 0.03 | 2.21(1.12-4.36)** |
| Age | ≤30 | 55 (63.9) | 0.97(0.59-1.59) | 0.49 | 1.29(0.69-2.39) |
|  | >30 | 31 (36.1) | 1 |  | 1 |
| Service year | 1-3 | 43 (50.0) | 1 |  | 1 |
|  | 3+ | 43 (50.0) | 1.38(0.86-2.22) | 0.18 | 1.13(0.64-2.01) |
| Training on COVID-19 | Yes | 27 (31.4) | 1.46(0.87-2.46) | 0.15 | 1.59(0.90-2.81) |
|  | No | 59 (68.6) | 1 |  |  |
| Profession | Medical doctor | 44 (51.2) | 1 |  | 1 |
|  | Specialist medical practitioners | 34 (39.5) | 1.58(0.95-2.61) | 0.07 | 1.60(0.86-2.99) |
|  | Resident physician | 8 (9.3) | 5.74(1.97-16.64)* | 0.001 | 7.10(2.30-21.95)** |
| Current working healthcare facility | Governmental | 74 (86.1) | 0.43(0.20-0.91)* | 0.02 | 0.48(0.22-1.08) |
|  | Private | 12 (13.9) | 1 |  | 1 |
| Department | OPD & Emergency | 27 (31.4) | 1 |  |  |
|  | Surgical Department & OR | 23 (26.7) | 0.76(0.41-1.41) | 0.39 |  |
|  | Medical Department | 14 (16.3) | 1.72(0.81-3.67) | 0.15 |  |
|  | Gynecology and Obstetrics Department | 6 (6.9) | 0.79(0.29-2.08) | 0.63 |  |
|  | Pediatric Department | 8 (9.3) | 1.55(0.62-3.88) | 0.38 |  |
|  | COVID-19 Center | 6 (6.9) | 3.05(0.97-9.54) | 0.06 |  |
|  | Intensive Care Unit (ICU) & others | 2 (2.3) | 0.74(0.15-3.54) | 0.71 |  |
| Availability of disinfectants in the workplace | Yes | 70 (81.4) | 2.96(1.08-3.54)* | 0.02 | 1.67(0.88-3.16) |
|  | No | 16 (18.6) | 1 |  | 1 |
| Are there any reminders about medical equipment disinfection in your working unit? (e.g. poster) | Yes | 21 (24.4) | 1.89 (1.06-3.37)* | 0.03 | 1.78(0.95-3.32) |
|  | No | 65 (75.6) | 1 |  | 1 |
| How often are you involved in direct contact with a known case of COVID-19 patient? | Daily | 20 (23.3) | 1.13(0.64-1.98) | 0.66 | 0.95(0.51-1.76) |
|  | Never/not yet/ | 66 (76.7) | 1 |  | 1 |

Hosmer-Lemeshow = 0.1631; Crude Odds Ratio: COR; Adjusted Odds Ratio: AOR; \*p-value < 0.05 (Crude); \*\*p-value < 0.05 (Adjusted)

## DISCUSSION

In order to scrutinize on the stethoscope and non-infrared thermometer disinfections, an online survey was conducted. This is the pivotal study about stethoscopes and non-infrared thermometer disinfections executed among the physicians in Ethiopia, and one of the few in the age of COVID-19 pandemic. The present study also dispensed insights about factors associated with disinfection after each use on patients. Overall, less than a fifth (13.9%) of the physicians disinfect their stethoscopes after every use, as needed; and a quarter (25.59%) of the respondents uttered that they have never disinfected their stethoscopes. This trend is similar to the antecedent studies done in different countries [4,8,36-38]. Despite empirical observation denoted the use of sanitizers and alcohol-based solutions in disinfecting non-critical medical equipments and other contaminated surfaces, and this had been ubiquitous in every corner of the globe. The present disturbingly low disinfection practice among physicians raises a concern in this unprecedented global crisis of coronavirus disease 2019 (COVID-19). As the person-to-person transmission of novel coronavirus has been outlined in the hospital settings [39], consistent and correct stethoscope disinfection should be observed after each use on patients [5,40], Upholding to the core values of nonmaleficence, physicians should be more diligent in providing safe domains for their patients.

To our cognizance, no specific guidelines solely on stethoscopes disinfection are available to date. The novelty of this pandemic – along with its uncertainties – makes it critical for health authorities to develop appropriate disinfection regulations to keep the safety for both patients and clinicians in the countenance of the current COVID-19 pandemic. The oft-cited Healthcare Infection Control Practices Advisory Committee (HICPAC) and Centers for Disease Control and Prevention (CDC) guidelines, advocating for non-critical medical equipment, such as stethoscopes, should be disinfected once daily or weekly [29]. The above standards might not be reflective on the current danger that a contaminated stethoscope may feasibly jeopardize patients’ and physicians’ safety – given the survival potential of coronavirus to thrive on different surfaces for an extended period of time [32]. It has been reported that SARS-CoV-2 can survive on steel and plastic surfaces for 72 hours or more [41]. Additionally, stethoscopes have high usance for assessment of COVID-19 patients [42, 43], and should be disinfected following each usage.

During times of increased disquietudes, technological advancements in the area of single-use aseptic diaphragm barriers provide a promising result for stethoscope hygiene [44]. Messina et al also propounded the utilization of pocket-held UV-LED devices which would be attached on the stethoscope diaphragms which will provide automatic disinfection of the stethoscope membranes [45]. On the downside, Alali et al connoted that despite the use of alcohol-based cleaner for stethoscope hygiene, physicians were ill-fated in eradicating all pathogenic microorganisms [46].

Often times, dialogues surrounding disinfections tend to focus only on critical equipment used for invasive procedures, like gastrointestinal endoscopes. However, essential medical equipment is easily neglected – such as non-infrared thermometers – but these can be prime sources of infection transmissions. In an early report, thermometers are linked to the outbreak of hospital-acquired infections [47, 48]. Until today, in most resource limited-settings, surveillance for body temperatures using non-contact digital thermometers are still shortly supplied, and the use of non-infrared thermometers are withal customary – often assumed sanitary, unless visibly soiled. Interestingly, our study corroborated that only a fifth of the physicians disinfected thermometers after every use, and a significant number of them (40.53%) never disinfected their thermometers at all. This finding has paramount implications in combating COVID-19, as contamination of routinely used devices in the healthcare settings are the possible sources of infections [32, 33]. A systematic review on the relationship between shared patient care items and healthcare-associated infections manifested that potential pathogens and multiple resistant organisms present on noninvasive portable clinical items (NPIs) in routine, non-outbreak conditions – and in a variety of settings affirms the need to improve NPIs decontamination practices [49].

In multivariable analyses, stethoscope disinfections after every use were significantly associated with the availability of the disinfectants in the workplaces. Previous studies also identified deficiencies on access to disinfection materials as potential barriers to stethoscope disinfections after every use [8]. In resource-limited settings, physicians often rely on clinical appraisals using the stethoscope, particularly the assessment of respiratory dysfunctions in COVID-19 positive patients [42], and eventually, these can cause the contagion of physicians and patients. Considering the communicability of the virus, the use of ultrasound is now recommended by Buonsenso et al as essential in the safe management of the COVID-19 pandemic; après it can allow the concomitant execution of clinical examination and lung imaging at the bedside by the same doctor [43]. However, in resource-limited settings, this may not be applicable. It is, therefore, of utmost importance that ensuring the availability of disinfectant solutions may precede a positive influence on disinfection compliance. Patel et al also argued that, at this point, it is difficult to answer if the stethoscope is a necessary tool or an unnecessary evil during COVID-19 pandemic, but removing it altogether from the care of patients with COVID-19 does not seem practical. And suggested basic guiding principles such as use of personal stethoscopes, use disposable isolation stethoscope and cleaning of stethoscope after patient auscultation with predefined stethoscope cleaning protocols [50].

Furthermore, providing trainings are beneficial, and thus significantly increases the odds of disinfection after every use – as confirmed from this study. The present study identified that the odds of stethoscope disinfection after every use were significantly reduced among those who reported the use of shared stethoscopes. In our point of view, sharing stethoscopes are detrimental, and may pose risks to physicians. Similar conclusions about the sharing of personal items have been described by WHO [34]. The researchers also believe that, if unavoidably stethoscopes are shared, it is paramount to disinfect the stethoscopes meticulously. Lastly, another fact but not of utmost significance, access to disinfectants in the workplaces (AOR 1.67; 95%CI: 0.88-3.16) increase the odds of non-infrared thermometer disinfections after ever use. In contrast, studies reported a positive correlation between availability of instructive posters and acceptable decontamination practice among healthcare workers [51-52].

### Limitations

This study has a number of limitations. First, recall bias may have affected our results. Second, external validation survey responses regarding direct observation on the physician’s disinfection practices were not executed. Third, the potential for social desirability biases among survey respondents are present, although the low reported stethoscope and non-infrared thermometer disinfections after ever use suggest that responses may not reflect the physicians’ actual behaviors. Fourth, the cross-sectional nature of the survey does not allow the cause-and-effect relationship between the dependent and independent variables. Fifth, the sample size was conducted into account the proportion of stethoscope disinfection practice but not for the associations of this disinfection practice, so in some cases, the power of the sample may not sufficient. And finally, this study should be generalizable for the physicians working in the healthcare facilities of this country.

## CONCLUSIONS

In summary, less than a fifth of the physicians disinfected their stethoscopes after every use, and a significant number of respondents reported that they have never disinfected their non-infrared thermometers. This study has reflected disturbing inadequate disinfection practices among physicians, whereby patient safety is of germane priority in the era of COVID-19. In this regard, physicians should be exigent and more vigilant in disinfecting these commonly used medical devices. Furthermore, this study provides empirical evidence on the association between the availability of disinfectants in the working units, and stethoscope disinfection after every use. With respect, measures should be taken by health authorities in upraising the current practices of disinfection through implementations of simple interventions, such as provisions of training and securing constant and available disinfection supplies.

## Data Availability

Availability of all data referred to in the manuscript is available from the corresponding author,with reasonable request.

## Appendices

**Table A.1: Physicians’ Non-Infrared Thermometer Disinfection**

